# Burden of Syphilis and STI Co-infections in Ghanaian Pregnant Women: Implications for Antenatal Screening Policy

**DOI:** 10.64898/2026.04.28.26351916

**Authors:** Anthony Zunuo Dongdem, Jennifer Sarpong, Emelia Narkie Sackitey, Kpedzi Evelyn, Ninyang Akosua Angela, Eric Yiadom Boakye, Sussan Akorfa Dzorgbedor, Eyram Kuma Hanu

## Abstract

**Background:** Sexually transmitted infections (STIs) pose major risks in pregnancy, including stillbirth, preterm delivery, and congenital infections. Globally, pooled antenatal prevalence estimates are HIV 2.9%, HBV 4.8%, HCV 1.0%, and syphilis 0.8%, but burdens are higher in low-income countries, reaching HIV 5.2%, HBV 6.6%, HCV 2.7%, and syphilis 3.3% (). In Ghana, reported rates are variable: HBV 4–10%, HCV 0.8–12%, HIV 1–3%, and syphilis 0.4– 3.6%. Despite a national policy mandating integrated antenatal screening, evidence on the prevalence and co-infection patterns among pregnant women remains fragmented. This study aimed to determine the prevalence of HIV, HBV, HCV, and syphilis, co-infections, and associated determinants among Ghanaian pregnant women.

**Methods:** A cross-sectional study was conducted using secondary data from 1,316 pregnant women attending antenatal care across four municipalities in Ghana (2023–2024). Prevalence, co-infection patterns, and risk factors were assessed using descriptive statistics, logistic regression, and kappa agreement tests.

**Results:** The median age of participants was 28 years (IQR: 23–33). Syphilis was most prevalent (10.5%), followed by HBV (4.0%), HIV (2.5%), and HCV (1.9%). Marked geographic disparities were observed, with syphilis prevalence ranging from 0.8% in Afigya Kwabre to 38.9% in Cape Coast. Co-infections were common: 26.6% of HBV-positive women also had syphilis (κ = 0.348, p < 0.001), and 16.6% of HIV-positive women had syphilis (κ = 0.237, p < 0.001). Predictors of syphilis included urban residence (aOR: 4.79; 95% CI: 2.99– 7.69), multiparity (aOR: 3.08; 95% CI: 1.92–4.96), and early gestational age.

**Conclusion:** The high burden of syphilis and frequent co-infections among Ghanaian pregnant women reveal critical gaps in implementing integrated antenatal screening. Despite national policy mandates, inconsistent practice leaves mothers and infants vulnerable to preventable complications. Strengthening compliance with comprehensive STI screening while tailoring interventions to high-risk groups is essential to reducing adverse maternal and neonatal outcomes.

## Introduction

Sexually transmitted infections (STIs) constitute a substantial global public health burden and are caused by viruses, bacteria, and parasites ^1^. Transmission occurs primarily through sexual contact and may also be vertical, from mother to child during pregnancy, delivery, or postnatally. Blood-borne infections such as HIV, hepatitis B virus (HBV), and hepatitis C virus (HCV) pose serious lifelong risks, whereas syphilis, chlamydia, and gonorrhoea are generally curable if detected and treated early. Pregnant women are particularly vulnerable due to immunologic changes in pregnancy, and untreated maternal infections can result in stillbirth, preterm delivery, low birth weight, neonatal death, congenital infections, and increased susceptibility to other pathogens ^2^.

Globally, more than one million curable STIs are acquired daily, with major consequences for maternal and neonatal health ^3^. A pooled global prevalence of blood-borne STIs among pregnant women has been estimated at HIV 2.9%, HBV 4.8%, HCV 1.0%, and syphilis 0.8%, with substantially higher burdens reported in low-income countries: HIV 5.2%, HBV 6.6%, HCV 2.7%, and syphilis 3.3% ^4^.

Across Africa, prevalence is high but heterogeneous. In Ethiopia, an overall STI prevalence of 15.2% was reported among pregnant women. In Lesotho, prevalence was 12.2% with HIV as high as 29.0% ^5^, while Tanzania recorded HIV 5.9%, HBV 4.7%, and syphilis 1.4% ^6^. In West

Africa, syphilis prevalence reached 6.8% in The Gambia and 1.8% in Nigeria, with HBV ranging between 4% and 10% ^7^.

In Ghana, prevalence estimates are variable across studies and regions. ^8^ reported HBV (10.0%) and HCV (12.0%); ^9^ observed an overall STI prevalence of 11.4% with HBV 9.8% and 0.8% each for HCV, HIV, and syphilis. Other studies reported lower prevalence, including HBV 4.2%, HIV 1.0%, and syphilis 0.4% ^10^, and syphilis 3.6% ^11^.

Co-infections further complicate the burden. A systematic review estimated co-infection rates ranging from 7.4% to 37.7%, with the highest levels in sub-Saharan Africa ^12^. In Botswana, HIV–syphilis co-infection reached 37.7%, with stillbirth and low birth weight more frequent among co-infected mothers ^13^. In Ghana, ^8^ documented HIV–HBV and HIV–HCV co-infections at 14.9% and 4.1%, respectively, with associated adverse pregnancy outcomes.

Ghanaian expectant mothers’ risk of STIs is shaped by behavioural factors such as unprotected sex and multiple partners, biological vulnerabilities from pregnancy-related immune changes, and social determinants including poverty and gender inequality. These factors heighten infection risk and contribute to adverse pregnancy outcomes.

Despite ongoing prevention and control efforts, progress in Ghana has been constrained by gaps in health education, routine screening, and treatment services, as well as challenges in implementing integrated antenatal screening policies. A major limitation is the lack of comprehensive and up-to-date regional and sub-regional data on STI prevalence and co-infections. This gap undermines effective surveillance, weakens clinical decision-making, and hinders targeted public health interventions. Addressing this is critical to achieving SDG 3.1 on reducing maternal mortality and SDG 3.3 on ending AIDS, controlling hepatitis, and eliminating syphilis as public health threats.

This study therefore seeks to quantify the prevalence of key blood-borne STIs (HIV, HBV, HCV, and syphilis), examine co-infection patterns, and identify sociodemographic determinants among pregnant women attending antenatal clinics in Ghana.

## 2. Materials and Methods

### Study design

A cross-sectional study design was employed to review 1-year secondary data of ANC attendees with STIs from 2023 to 2024 in selected municipalities in Ghana: Kpandai, Cape Coast, Ayawaso West, and Afigya Kwabre.

### Study site

The study was conducted in four selected municipalities in Ghana. Cape Coast Metropolitan had a total population of 169,894 including 93,619 females and 89,017 males with approximately 80,000 reproductive age women. Ayawaso West Municipal’s had a population of 75,303 with 38,614 males and 36,689 females with a population of approximately 36,689 reproductive age women. Kpandai district had a population of approximately 126,213, including a significant number of women of reproductive age. Afigya Kwabre had a population of 200,000, with about 100,000 women of reproductive age (GSS,2021). Key facilities included 37 Military Hospital, Ridge Hospital, Korle-Bu Teaching Hospital, Cape Coast Teaching Hospital, Regional Hospital, District hospitals, health centres, CHPS compounds and various polyclinics.

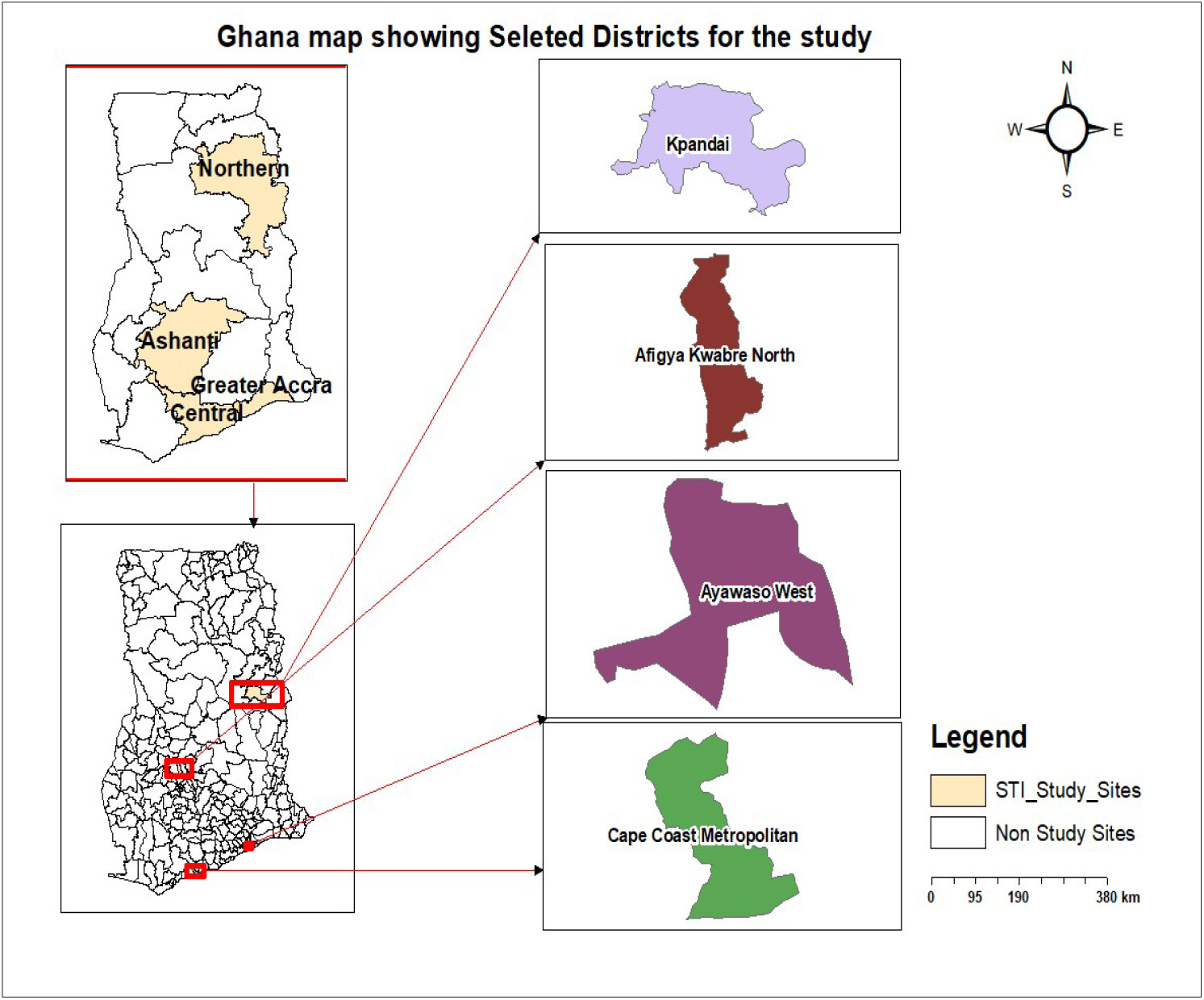

#### Study Population

The study population consisted of pregnant women who attended antenatal clinics from 2023to 2024in selected districts.

##### Sample and Sampling strategy

The study utilized a purposive sampling approach to select health facilities and a census method to include all eligible 1,316 pregnant women who attended the antenatal clinic within the study period. Health facilities were deliberately chosen based on three key criteria: provision of antenatal care (ANC) services, routine STI testing for pregnant women as part of ANC, and availability of complete STI records from 2023 to 2024. This targeted facility selection ensured the inclusion of relevant data sources that could provide comprehensive information about STI prevalence among pregnant women attending ANC clinics.

##### Inclusion and Exclusion

The study included STI test data from all pregnant women attending antenatal care in the selected district hospitals while women with incomplete medical/STI test records, untested individuals, and on repeated visits were excluded.

##### Data collection

STI data (2023-2024) were collected from six purposively selected Ghanaian health facilities across the selected districts, chosen for high patient volume and regional diversity. Using stratified sampling by district, facility type, and case load, records were systematically randomized within each stratum via computer-generated selection to ensure representative, unbiased data reflecting regional STI trends.

#### Data management and statistical analysis

Data were compiled and cleaned in Excel, and analysed using STATA v17.0. Descriptive statistics: frequencies, means, proportions summarized the dataset, while inferential analyses included logistic regression and Kappa statistics. The Kappa statistic (κ) quantified agreement between observed and expected co-infection rates, with interpretation thresholds: κ<0.20 (slight), 0.21–0.40 (fair), 0.41–0.60 (moderate), 0.61–0.80 (substantial), and >0.80 (almost perfect agreement).

#### Ethical consideration

Ethical approval for this study was obtained from the University of Health and Allied Sciences Research Ethics Committee (Approval No. UHAS-REC B.10 [52]24-25). Permission to access and use the data was also granted by the management of Ho Teaching Hospital and Hohoe Regional Hospital.

The study was conducted in accordance with the principles of the Declaration of Helsinki. As this study involved retrospective analysis of anonymized secondary data from blood donor registers, a waiver of informed consent was granted by the ethics committee. No personal identifiers were collected, and all data were handled with strict confidentiality.

## 3. Results

### Socio-Demographic Characteristics and Prevalence of HBV, HCV, HIV, and STIs (Syphilis) among mothers attending Antenatal Clinic from 2023 to 2024 in the selected districts (N = 1316)

Table 3.1 summarizes the socio-demographic characteristics of the study participants and the prevalence of HBV, HCV, HIV, and syphilis. A total of 1,316 pregnant women were included, with a median age of 28 years (IQR: 23–33). The majority were aged 26–30 years (353/1,316; 26.8%), in their second trimester (525/1,316; 39.9%), and multiparous (525/1,316; 39.9%). Most participants resided in rural areas (867/1,316; 65.9%), and secondary education was the most common educational level (491/1,316; 37.3%).

**Table 3.1:**
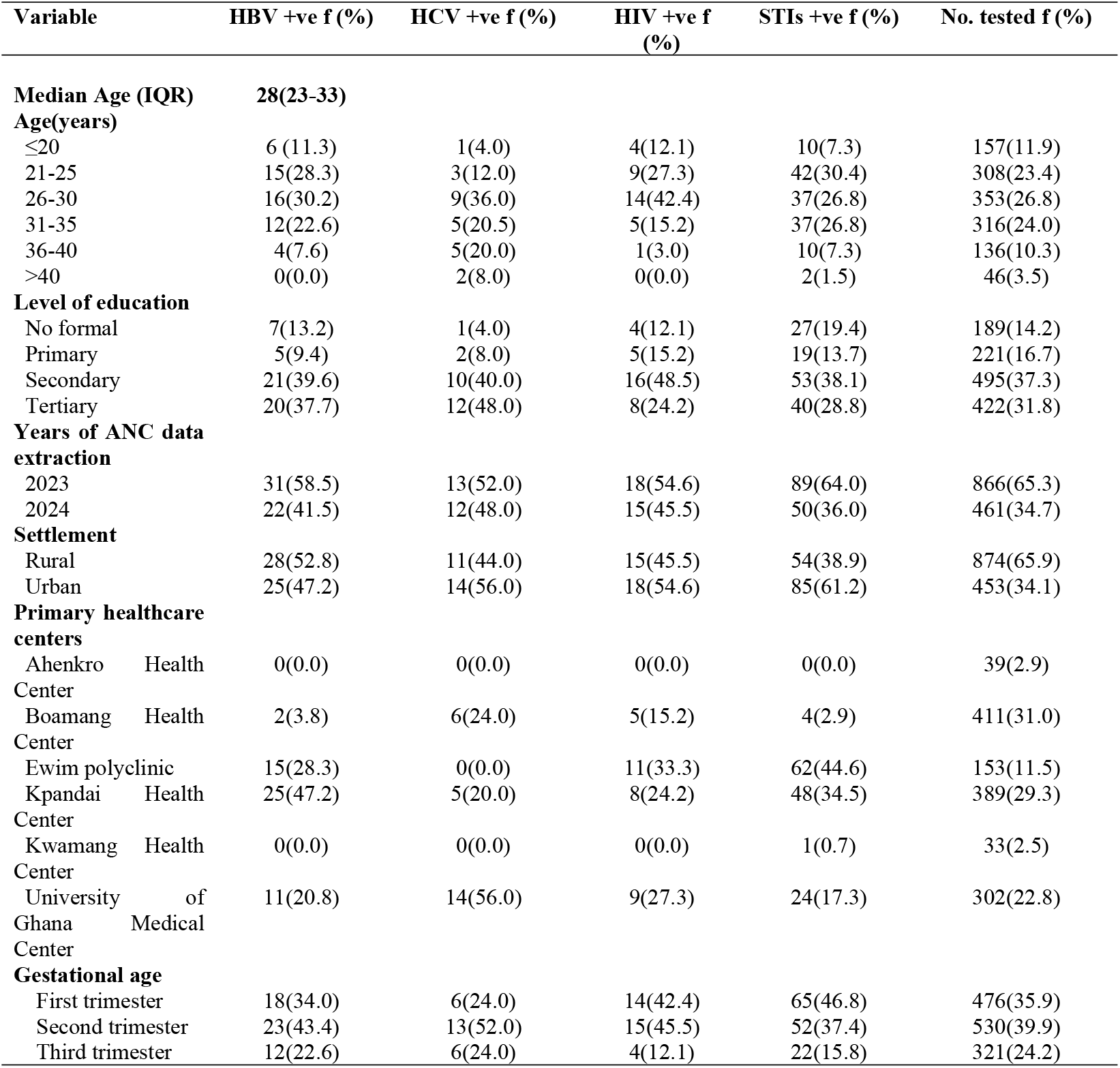
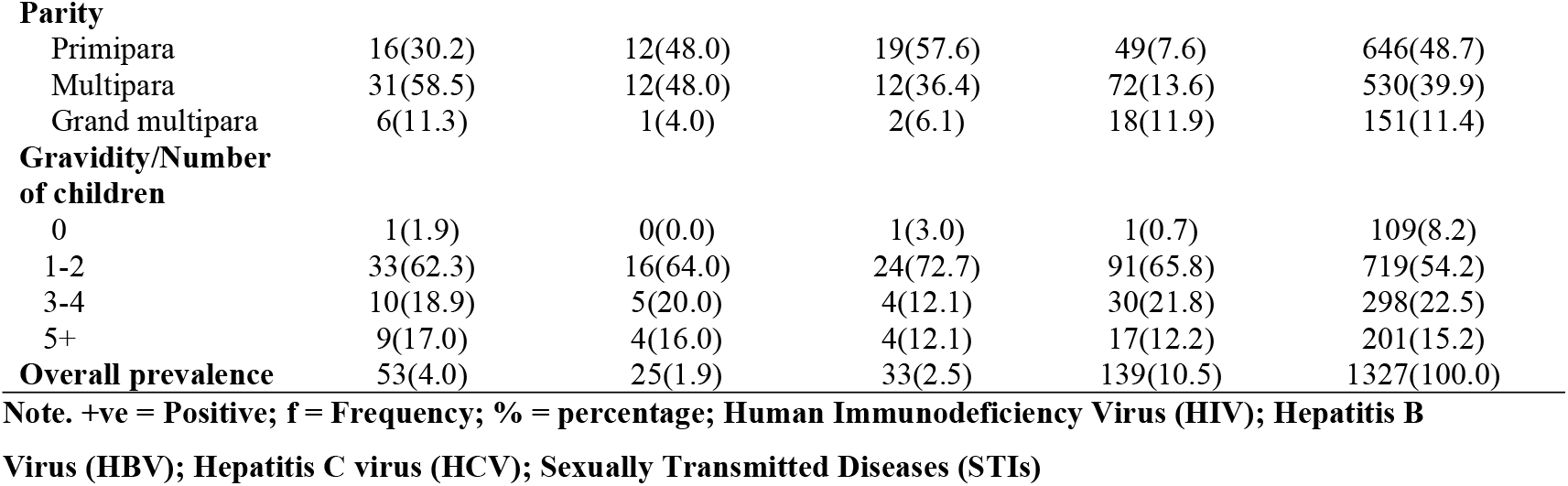
Socio-Demographic Characteristics and Prevalence of HBV, HCV, HIV, and STIs (Syphilis) among mothers attending Antenatal Clinic in selected districts from 2023 to 2024.

The overall prevalence of hepatitis B virus (HBV) was 4.0% (53/1,316), hepatitis C virus (HCV) 1.9% (25/1,316), HIV 2.5% (33/1,316), and syphilis 10.5% (139/1,316).

HBV infection was most frequent among women aged 26–30 years (16/53, 30.2%), in their second trimester (23/53, 43.4%), multiparous (31/53, 58.5%), rural residents (28/53, 52.8%) and had attained secondary education (21/53, 39.6%). The highest number of HBV cases were recorded at Kpandai Health Centre (25/53, 47.2%).

For HCV, prevalence was highest among women aged 26–30 years (9/25, 36.0%). Most HCV-positive women were in their second trimester (13/25, 52.0%), multiparous (12/25, 48.0%), urban residents (14/25, 56.0%), and with tertiary education (12/25, 48.0%). Over half of the HCV cases were reported at the University of Ghana Medical Centre (14/25, 56.0%).

HIV prevalence peaked in the 26–30 age group (14/33, 42.4%), with most cases among first-trimester women (14/33, 42.4%), primiparous (19/33, 57.6%) and attained secondary education (16/33, 48.5%). Urban residents accounted for more than half of HIV cases (18/33, 54.6%). The highest site-specific prevalence was at Ewim Polyclinic (11/33, 33.3%).

Syphilis was the most common infection overall, with 10.5% prevalence (139/1,316). The highest burden was observed among women aged 21–25 years (42/139, 30.4%), those in their first trimester (65/139, 46.8%), primiparous mothers (49/139, 35.3%), with secondary education (53/139, 38.1%), and urban residents (85/139, 61.2%). Syphilis cases were concentrated at Ewim Polyclinic (62/139, 44.6%) and Kpandai Health Centre (48/139, 34.5%).

### Prevalence of HBV, HCV, HIV, and STIs (Syphilis) among mothers attending Antenatal Clinic from 2023 to 2024 across the selected Districts (N=1316)

Table 3.2 shows that prevalence varied markedly by district. Cape Coast recorded the highest syphilis (63/162, 38.9%), HBV (15/162, 9.3%) and HIV (11/162, 6.8%), whereas Afigya Kwabre reported the lowest prevalence for all infections (syphilis 4/475, 0.8%; HBV 2/475, 0.42%). Ayawaso West recorded the highest HCV prevalence (14/303, 4.6%), while Kpandai showed moderate levels of HBV (25/387, 6.5%) and syphilis (48/387, 12.4%). These inter-district differences were statistically significant (HBV: p < 0.001; syphilis: p < 0.001; HCV: p = 0.002; HIV: p = 0.001).

**Table 3.2:**
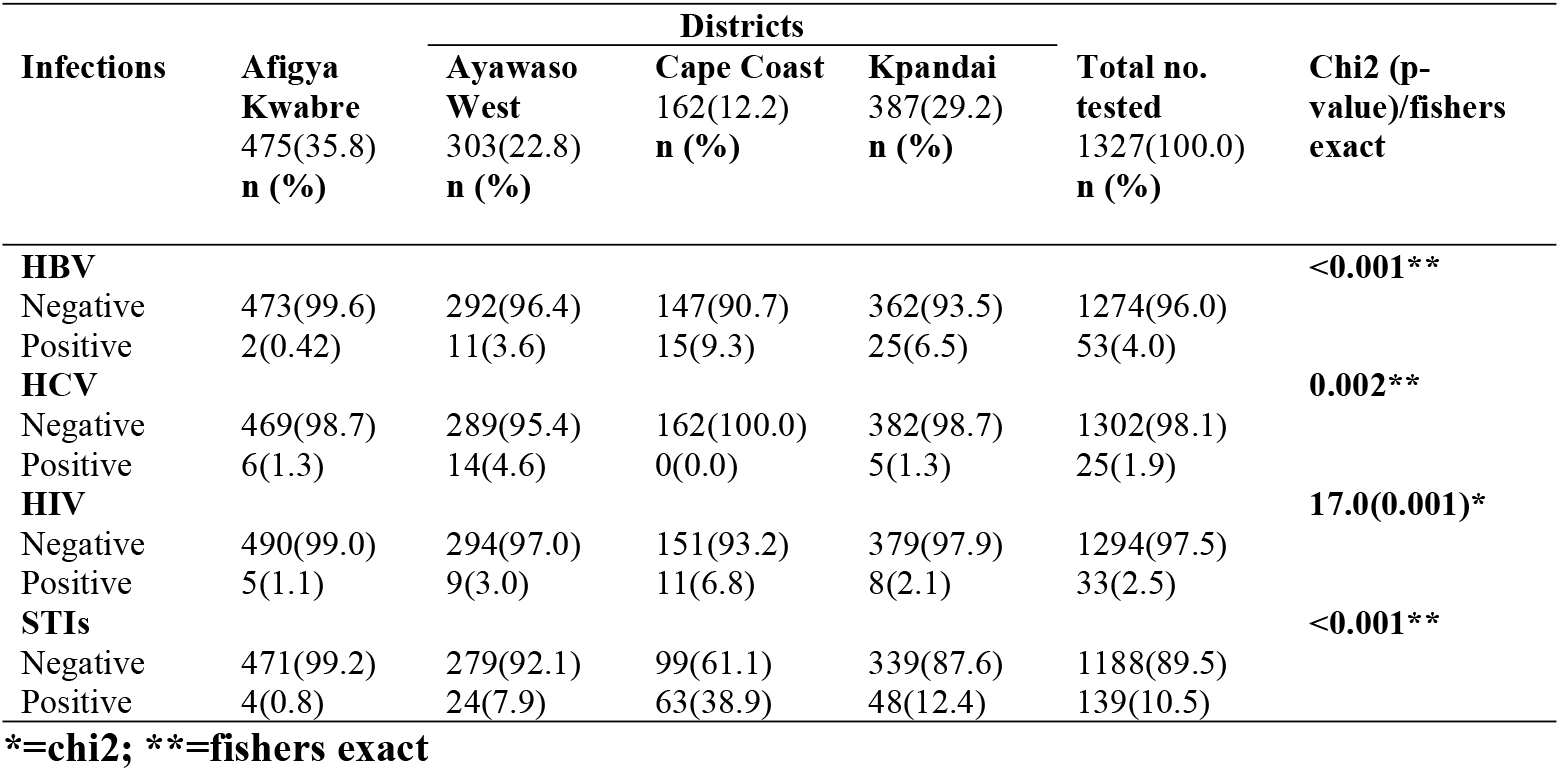
Prevalence of HBV, HCV, HIV, and STIs (Syphilis) among mothers attending Antenatal Clinic from 2023 to 2024 in four selected Districts (N=1316)

### Co-infections prevalent rate of HBV, HCV, HIV, and STIs (Syphilis) among mothers attending Antenatal Clinic in the selected districts (N = 1316)

**Table 3.3 shows** the prevalence of co-infections among the study participants. HIV-positive mothers were more likely to also have HBV [5/53 (9.4%)] and syphilis [23/139 (16.6%)], with slight (κ = 0.088, p = 0.001) and fair (κ = 0.237, p < 0.001) agreement, respectively. HIV–HCV co-infection was rare [1/25 (4.0%)] and not statistically significant (κ = 0.013, p = 0.312). For HBV, co-infection with HCV was observed in 3/25 (12.0%) cases (κ = 0.053, p = 0.020), while HBV–syphilis co-infection was more common [37/139 (26.6%)] with a strong association (κ = 0.348, p < 0.001). HCV–syphilis co-infection was very rare [3/139 (2.2%)] and not significant (κ = 0.005, p = 0.401). Overall, meaningful associations were observed between HBV–syphilis and HIV–syphilis, whereas HCV co-infections were infrequent.

**Table 3.3:**
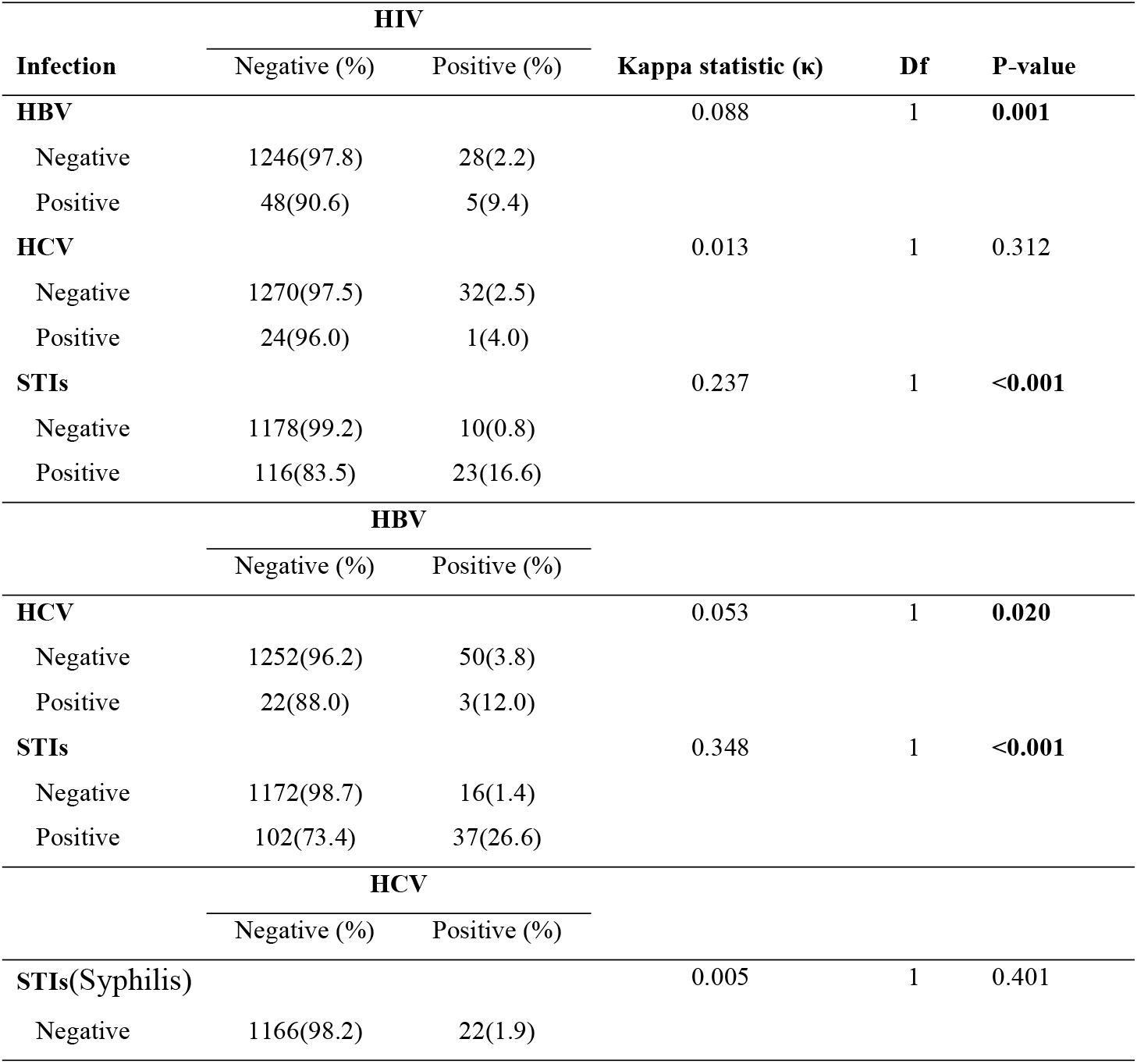
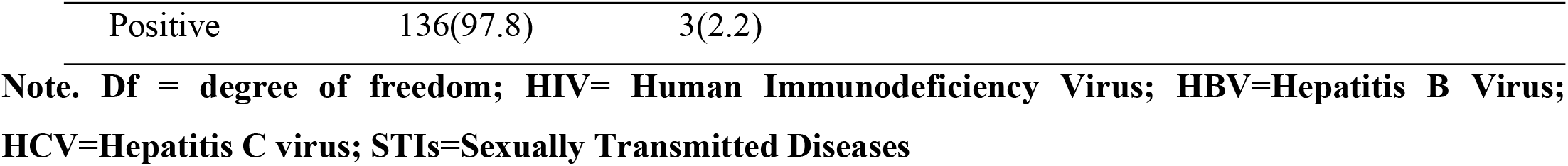
Co-infections prevalent rate of HBV, HCV, HIV, and STIs (Syphilis) among mothers attending Antenatal Clinic in the selected districts (N = 1316)

### Sociodemographic factors associated with STI prevalence among mothers attending Antenatal Clinic in the selected Districts

**Table 3.4** shows the results of the bivariate and multivariate logistic regression analyses of sociodemographic factors associated with syphilis prevalence among mothers attending antenatal clinics. Multivariate logistic regression identified age, settlement, parity, gravidity, and gestational age as significant predictors of syphilis prevalence. Mothers aged 36–40 years had significantly lower odds of infection compared to those aged ≤20 years (aOR: 0.29; 95% CI: 0.10–0.84; p = 0.023), while those older than 40 years also had reduced odds (aOR: 0.18; 95% CI: 0.03–1.00; p = 0.050). Urban residence was strongly associated with increased syphilis prevalence compared to rural residence (aOR: 4.79; 95% CI: 2.99–7.69; p < 0.001). Gestational age was inversely associated with syphilis, with women in their second trimester having lower odds compared to those in the first trimester (aOR: 0.64; 95% CI: 0.42–0.97; p = 0.034), and those in the third trimester showing even greater reductions (aOR: 0.34; 95% CI: 0.20–0.59; p < 0.001). Parity was also strongly related to syphilis prevalence, as multiparous women had higher odds compared to primiparous mothers (aOR: 3.08; 95% CI: 1.92–4.96; p < 0.001), and grand multiparous women had the highest risk (aOR: 4.27; 95% CI: 1.80–10.1; p = 0.001). Similarly, gravidity showed significant associations, with women who had five or more children exhibiting markedly higher odds of syphilis compared to those with no children (aOR: 8.95; 95% CI: 1.01–79.6; p = 0.049). Educational level was not independently associated with syphilis prevalence after adjustment.

**Table 3.4:** Bivariate and Multivariate logistic regression analyses of Sociodemographic factors associated with STI (Syphilis) prevalence among mothers attending Antenatal Clinic in the Selected Districts.

| Variable | Prevalence of STI (Syphilis) |  | cOR (CI 95%), P-value | aOR (CI 95%), P-value |
| --- | --- | --- | --- | --- |
|  | Negative (n=89.5) | Positive (n=10.5) |  |  |
|  | n (%) | n (%) |  |  |
| **Age(years)** |  |  |  |  |
| ≤20 | 147(93.6) | 10(6.4) | Ref | Ref |
| 21-25 | 266(86.4) | 42(13.6) | **2.32(1.13-4.76), 0.022** | 1.28(0.59-2.76), 0.536 |
| 26-30 | 316(89.5) | 37(10.5) | 1.72(0.83-3.56), 0.142 | 0.55(0.24-1.25), 0.154 |
| 31-35 | 279(88.3) | 37(11.7) | 1.95(0.94-4.03), 0.072 | 0.48(0.20-1.13), 0.092 |
| 36-40 | 126(92.7) | 10(7.4) | 1.17(0.47-2.89), 0.739 | **0.29(0.10-0.84), 0.023** |
| >40 | 44(95.7) | 2(4.4) | 0.67(0.14-3.16), 0.611 | 0.18(0.03-1.0), 0.050 |
| **Level of education** |  |  |  |  |
| No formal | 162(85.7) | 27(14.3) | Ref |  |
| Primary | 202(91.4) | 19(8.6) | 0.56(0.30-1.05), 0.072 |  |
| Secondary | 442(89.3) | 53(10.7) | 0.72(0.44-1.18), 0.194 |  |
| Tertiary | 382(90.5) | 40(9.5) | 0.63(0.37-1.06), 0.081 |  |
| **Settlement** |  |  |  |  |
| Rural | 820(93.8) | 54(6.2) | Ref | Ref |
| Urban | 368(81.2) | 85(18.8) | **3.51(2.44-5.04), <0.001** | **4.79(2.99-7.69), <0.001** |
| **Gestational age** |  |  |  |  |
| First trimester | 411(86.3) | 65(13.7) | Ref | Ref |
| Second trimester | 478(90.2) | 52(9.8) | 0.69(0.47-1.01), 0.059 | **0.64(0.42-0.97), 0.034** |
| Third trimester | 299(93.2) | 22(6.9) | **0.47(0.28-0.77), 0.003** | **0.34(0.20-0.59), <0.001** |
| **Parity** |  |  |  |  |
| Primipara | 597(92.4) | 49(7.6) | Ref | Ref |
| Multipara | 458(86.4) | 72(13.6) | **1.92(1.31-2.81), 0.001** | **3.08(1.92-4.96), <0.001** |
| Grand multipara | 133(88.1) | 18(11.9) | 1.65(0.93-2.92), 0.087 | **4.27(1.80-10.1), 0.001** |
| **Gravidity/Number of pregnancies** |  |  |  |  |
| 0 | 108(99.1) | 1(0.9) | Ref | Ref |
| 1-2 | 628(87.3) | 91(12.7) | **15.7(2.16-113.5), 0.007** | 7.49(0.99-56.7), 0.051 |
| 3-4 | 268(89.9) | 30(10.1) | **12.1(1.63-89.8), 0.015** | 6.42(0.79-52.0), 0.081 |
| 5+ | 184(91.5) | 17(8.5) | **9.98(1.31-76.0), 0.026** | **8.95(1.01-79.6), 0.049** |

## 4. DISCUSSION

Sexually transmitted infections (STIs) among pregnant women in Ghana remain a significant public health concern, posing risks to maternal and neonatal health. Limited regional and sub-regional data on prevalence and co-infections hinder effective surveillance, clinical management, and public health interventions. This study assessed the prevalence and co-infection rates of HIV, hepatitis B, hepatitis C, and syphilis among pregnant women attending antenatal clinics across selected Ghanaian districts from 2023 to 2024 and examined associated sociodemographic factors. The findings provide current evidence to inform targeted antenatal interventions and guide policy reforms aligned with SDG 3.1 on reducing maternal mortality and SDG 3.3 on combating HIV, controlling hepatitis, and eliminating syphilis, while contributing to the literature on maternal health and infectious diseases in Ghana.

### Prevalence of Individual STIs

This study revealed a heterogeneous burden of sexually transmitted infections (STIs) among pregnant women attending antenatal clinics in Ghana, with notable infection-specific prevalence rates: syphilis showed the highest prevalence at 10.5%, followed by hepatitis B virus (HBV) at 4.0%, human immunodeficiency virus (HIV) at 2.5%, and hepatitis C virus (HCV) at 1.9%. The markedly higher prevalence of syphilis contrasts with the relatively lower rates of HBV, HIV, and HCV, highlighting ongoing maternal health challenges in Ghana ^14^.

Syphilis prevalence at 10.5% indicates a persistently high burden among pregnant women in Ghana, far exceeding recent regional estimates of 0.8% to 4.1% ^15, 9^. West African and African averages of 2–6%, and the global antenatal rate of 1.6% ^1, 4^. This discrepancy may arise from variations in screening coverage, diagnostic practices, and treatment access. The findings signal ongoing transmission due to high economic and tourism activities, sociocultural practices, risky sexual behaviours and critical gaps in antenatal screening and treatment integration. The disproportionately high burden among younger women aged 21–25 and primiparous mothers points to the need for prioritized interventions focused on early pregnancy screening and targeted sexual health education ^14^.

The HBV prevalence of 4.0% indicates moderate endemicity, reflecting ongoing progress through effective vaccination programs, including birth dose and infant immunizations. This aligns with antenatal prevalence ranges of 3–8% nationally and across West Africa ^9, 7, 16^ and global prevalence of 3–4% ^17^. Higher prevalence in rural areas, multiparous women, ages 26– 30, and during the second trimester likely reflects lower vaccination coverage, cumulative exposure, and screening timing differences. These factors highlight the need for targeted vaccination, education, and improved healthcare access ^18, 19, 9^.

A moderate HIV prevalence of 2.5% reflects progress from widespread antenatal testing, counselling, antiretroviral therapy, and the national PMTCT program. This prevalence is slightly higher than recent national antenatal surveillance estimates of approximately 1.66% by GHS ^20^, which reflects a decline from earlier prevalence levels above 3% over the past decade ^21^. It is slightly lower than West African averages of 3–5%, but above the global antenatal estimate of 1% ^22, 1^. Higher rates in urban facilities and younger, primiparous women likely reflect biological vulnerability and structural drivers such as higher partner concurrency, increased mobility, and earlier ANC attendance that facilitates diagnosis ^23^. Despite progress, challenges persist, including urban–rural disparities in service access, retention in PMTCT programs, stigma, and low uptake of partner testing, which collectively hinder further gains. Strengthening targeted urban-focused prevention while addressing systemic barriers in PMTCT delivery will be critical to achieving sustained reductions in HIV prevalence and eliminating vertical transmission.

The low HCV prevalence of 1.9% reflects limited exposure to high-risk factors compared to other infections, consistent with national estimates in Ghana ranging from 1.5% to 4.6% ^24, 25^. Within West Africa, it aligns with generally low prevalence under 2% ^9, 26^, though it contrasts with a higher prevalence reported in Nigeria. The prevalence is slightly higher than the global antenatal rate of 1%. Urban and tertiary-educated women showed higher prevalence, likely due to exposure to medical procedures, intravenous drug use, and lifestyle factors. This highlights the need for urban-centred targeted screening and harm reduction programs.

Geographically, district-level analysis revealed Cape Coast as the highest-burden district for HBV (9.3%), HIV (6.8%), and syphilis (38.9%). This observation aligns with its profile as a densely populated urban and tourist hub characterised by high mobility and transient populations, factors known to intensify STI transmission. Similar regional variation across districts has been reported in Ghana ^25, 23^ likely reflecting uneven access to services, differences in screening coverage, and varied exposure risks. Cape Coast’s higher prevalence underscores both social and systemic factors. HCV prevalence was highest in Ayawaso West (4.6%), reflecting urban-specific risk exposures. Such variations underscore the need for tailored public health interventions and resource allocation based on local epidemiology ^25, 23^.

Overall, the study situates Ghana within global and continental epidemiological landscapes, evidencing progress in HIV and HBV control but highlighting the considerable gap in syphilis prevention. These findings underscore the need for mandatory antenatal screening for HIV, HBV, and syphilis, coupled with context-specific preventive measures and health education to reduce maternal and neonatal morbidity.

### Prevalence of STI Co-infections

This study highlighted notable co-infections among HIV, HBV, HCV, and syphilis, with the most significant associations observed for HIV–syphilis and HBV–syphilis, while HCV-related co-infections were rare and statistically negligible.

The higher prevalence of HIV–syphilis likely reflects shared sexual transmission and the facilitation of HIV acquisition by syphilitic ulcers, compounded by behavioural risks and limited antenatal screening. This finding contrasts with a Tanzanian study ^27^ but aligns with reports from Sub-Saharan Africa, where elevated co-infection rates persist. Globally, lower prevalence is reported in Asia and Europe due to stronger screening and treatment programs. These findings indicate ongoing STI burden and the need for strengthened antenatal interventions.

Similarly, the strong association observed between HBV and syphilis co-infection may reflect overlapping risk factors. This finding contrasts with reports from Ghana ^9^ and Ethiopia ^28^, where the prevalence of HBV–syphilis co-infection was markedly lower. While HBV can be transmitted sexually, vertically, or via blood exposure, syphilis is primarily sexual and vertical. The co-occurrence of these infections may therefore indicate shared sexual exposure, low vaccination coverage for HBV, or delayed syphilis diagnosis in antenatal settings. While HBV– syphilis co-infections are rarely highlighted in global studies, data from Asia and North America suggest much lower dual prevalence, possibly due to universal HBV vaccination and robust maternal screening systems. This underscores the need for integrated and mandatory HBV and syphilis screening in maternal clinics.

Although HIV–HBV co-infection was observed, the association was weaker (slight agreement), suggesting that while co-occurrence exists, it may be less clinically impactful in this population. Nevertheless, it highlights the need for continued attention to HBV screening and immunization among HIV-positive mothers. HCV-related co-infections, including HIV– HCV, HBV–HCV, and HCV–syphilis, were rare and mostly non-significant, reflecting differences in transmission dynamics. HCV is primarily bloodborne, with sexual transmission being relatively inefficient, which likely explains its low co-occurrence in this cohort.

These findings highlight the need to strengthen mandatory antenatal screening for HIV, HBV, and syphilis, supported by HBV vaccination and timely treatment to reduce maternal and neonatal risks. Preventive measures must be context-specific, with tailored health education addressing local behavioural and healthcare challenges.

### Sociodemographic Factors Associated with STI(Syphilis) Prevalence

This study found that syphilis prevalence among pregnant women was significantly influenced by age, settlement, gestational age, parity, and gravidity, while education was not independently associated.

Older maternal age was protective. Women aged 36–40 years had 71% lower odds (aOR: 0.29; 95% CI: 0.10–0.84; p = 0.023), and those over 40 had 82% lower odds (aOR: 0.18; 95% CI: 0.03–1.00; p = 0.050) compared with ≤20 years. This suggests that younger mothers are more vulnerable due to biological susceptibility, limited STI knowledge, and higher-risk behaviours. While similar protective effects have been documented in Ethiopia and Ghana ^28, 29^. Tanzania reported higher odds among older women ^30^, and Nigeria found no consistent pattern ^31^. These discrepancies may reflect differences in sexual debut, marital patterns, cumulative exposure, and access to testing. The implication is that interventions must prioritize adolescents and young mothers for early testing, treatment, and sexual health education.

Urban women had nearly five times higher odds of syphilis (aOR: 4.79; 95% CI: 2.99–7.69; p < 0.001) compared to rural mothers. This aligns with Ghanaian and other African urban studies that associate urbanization with increased STI risk due to high population density, mobility, and transactional sex ^23, 30^. Yet, Tanzanian national data reported higher rural prevalence ^30^, highlighting heterogeneity. Such differences may stem from sexual network structures, ANC coverage, and socioeconomic environments. In Ghana, rapid urbanization and gaps in partner treatment sustain transmission, implying a need for urban-focused strategies such as point-of-care testing, immediate treatment, and targeted education in dense ANC clinics.

Moreover, gestational age was inversely related to syphilis. Second-trimester women had 36% lower odds (aOR: 0.64; 95% CI: 0.42–0.97; p = 0.034), and third-trimester women had 66% lower odds (aOR: 0.34; 95% CI: 0.20–0.59; p < 0.001) than first-trimester mothers. This likely reflects early ANC detection and treatment, which reduces infection later in pregnancy. However, some West African studies found no consistent association, possibly due to varied booking times and testing protocols. Survivor bias may also play a role, as untreated early infections can result in miscarriage or stillbirth ^32^. These findings reinforce the importance of first-trimester screening and same-day treatment to prevent congenital syphilis and adverse pregnancy outcomes.

Finally, reproductive history showed a strong link with infection. Multiparous mothers had three times higher odds (aOR: 3.08; 95% CI: 1.92–4.96; p < 0.001), grand multiparous women had four times higher odds (aOR: 4.27; 95% CI: 1.80–10.1; p = 0.001), and women with gravidity ≥5 had nine times higher odds (aOR: 8.95; 95% CI: 1.01–79.6; p = 0.049). Similar patterns are reported in Ghana, Nigeria, Cameroon, and Tanzania ^33, 34, 30^, attributed to cumulative exposure and reduced uptake of preventive services. Still, differences across settings may reflect cultural norms, ANC utilization, and partner treatment coverage. For Ghana, repeated pregnancies may increase exposure to untreated partners and strain ANC services, implying the need for enhanced screening, counselling, and partner services tailored to multiparous and grand multiparous mothers.

Although education was not independently associated with syphilis after adjustment, higher educational attainment appeared to show some protective effect at the crude level. This suggests that education may still play a role indirectly, possibly through improved health-seeking behaviours, but its effect was attenuated by stronger predictors such as urban residence, parity, and gravidity.

The findings highlight how demographic, reproductive, and contextual factors intersect to shape syphilis risk in pregnancy. Addressing these drivers requires integrated antenatal and community interventions that combine early detection, effective treatment, and tailored prevention strategies.

## 5. LIMITATIONS

This study had some limitations. Its cross-sectional design made it difficult to determine causal relationships. Additionally, the analysis was restricted to selected healthcare facilities, which may limit representativeness at the district level and reduce generalizability. Finally, reliance on hospital-based data may not capture infections in the wider community, constraining the applicability of the findings to the general population. Future research should incorporate a broader range of facilities and community-based data to enhance representativeness and strengthen inference.

## 6. CONCLUSION

This study revealed a notably high prevalence of syphilis (10.5%) among pregnant women in Ghana, surpassing rates of HBV, HIV, and HCV. While descriptive trends were observed across all infections, only syphilis was examined inferentially, with younger maternal age, urban residence, early gestation, and high parity/gravidity identified as significant predictors. Although education was not independently associated, crude trends suggested a modest protective role through health-seeking behaviours. Importantly, despite Ghana’s national policy on integrated ANC screening, implementation remains inconsistent, with HIV testing often prioritized over HBV, HCV, and syphilis.

Strengthening maternal health therefore requires mandatory compliance with routine and integrated ANC screening, ensuring that all four infections are equally prioritized. Complementary measures include community health education for younger and urban women, engagement of local leaders to reduce stigma and encourage care-seeking, and expansion of sexual health education policies for adolescents. At the system level, community-based STI monitoring should be enhanced to detect outbreaks early and guide interventions.

The unexpectedly high syphilis burden uncovered in this study underscores the urgent need to elevate syphilis prevention and control as a maternal health priority in Ghana. Future studies should evaluate facility-level compliance with integrated ANC screening and its effectiveness in reducing adverse maternal and neonatal outcomes.

## Data Availability

N/A

## Acknowledgements

We are most grateful to the Directors of Health Services and their staff at the Afigya Kwabre North Health Directorate, Cape Coast Metropolitan Health Directorate, and Kpandi Health Directorate for granting us permission to use the data for this study. We are also deeply indebted to the district hospitals, health facilities, and antenatal care (ANC) units for their support during data collection. Finally, we sincerely appreciate the laboratory staff for their invaluable assistance in the data collection process.

## Authors Contribution

A.Z.D., J., E.N.S., K.E., and N.A.A. conceptualized the study. J.S.E., E.N.S., K.E., and N.A.A. were responsible for data collection. E.Y.B., S.A.D., and E.K.H. conducted the data analysis. E.Y.B., S.A.D., and E.K.H. drafted the manuscript, while A.Z.D. and E.K.H. critically reviewed it for important intellectual content. All authors reviewed, revised, and approved the final manuscript.

## Data Availability Statement

All relevant data are within the manuscript.

## Clinical Trial

Not applicable

## Funding

The authors received no specific funding for this work.

## Competing Interests

The authors have declared that no competing interests exist.

## Notes

### Competing Interest Statement

The authors have declared no competing interest.

### Funding Statement

The author(s) received no specific funding for this work.

### Author Declarations

Ethical approval for this study was obtained from the University of Health and Allied Sciences Research Ethics Committee (Approval No. UHAS-REC B.10 [52]24-25).

## REFERENCES

1. World Health Organization. Sexually transmitted infections (STIs). 2023. Available from: https://www.who.int/news-room/fact-sheets/detail/sexually-transmitted-infections-(stis)

2. Eskinder I. Global burden of sexually transmitted infections and their impact on pregnancy outcomes. 2023

3. World Health Organization. New report flags major increase in sexually transmitted infections amidst challenges in HIV and hepatitis. Geneva: WHO; 2024

4. Wu L, Huang S, Wu L. Epidemiology of sexually transmitted infections among pregnant women: a global meta-analysis. Lancet Glob Health. 2023;11(1):100–115

5. Xavier P, Magetse J, Monnapula-Mazabane P, Letsa L. Prevalence of syphilis and associated factors among pregnant women in Lesotho. Open Public Health J. 2024;16:e18749445255982

6. Mboya IB, Mremi IR, Majigo MV. Prevalence and factors associated with syphilis infection among pregnant women in Tanzania. BMC Pregnancy Childbirth. 2023;23:214

7. Isara AR, Baldeh I. Prevalence and risk factors of hepatitis B and syphilis co-infection among pregnant women. Afr Health Sci. 2021;21(2):450–456

8. Frempong K, Sagoe K, Doe E, Odame EA, Atsu BK. Co-infection of HIV and hepatitis B or C among pregnant women in Ghana: prevalence and risk factors. BMC Pregnancy Childbirth. 2019;19(1):123

9. Kuugbee ED, Abudukari M, Aryee PA, Abdul-Karim A. Seroprevalence and risk factors of sexually transmitted blood-borne infections among pregnant women in Northern Ghana. J Immunol Res. 2023;2023:3157202

10. Helegbe GK, Ofosu W, Baffoe P, Amoako YA. Prevalence of hepatitis B and C infections among pregnant women in selected Ghanaian hospitals. Ghana Med J. 2018;52(3):140– 146

11. Atupra SAA, Beyuo T, Lawrence ER. Knowledge about sexually transmitted infections among sexual and reproductive health clinic attendants in Ghana. Health Sci Investig J. 2021;2(2):230–237

12. Mendoza C, Oliva SM, Osorio A. Global and regional prevalence estimates of maternal syphilis and HIV co-infection: a systematic review. Int J Gynecol Obstet. 2018;141(2):123–130

13. Shava E, Moyo S, Zash R, Diseko M, Dintwa EN, Mupfumi L, et al. High rates of adverse birth outcomes in HIV and syphilis coinfected women in Botswana. J Acquir Immune Defic Syndr. 2019;81(5):E135–E140

14. Boachie J, Akakpo PK, Nimo P, Ofori-Asenso R. Syphilis infection in pregnancy: prevalence, risk factors, and impact on birth outcomes in Ghana. BMC Pregnancy Childbirth. 2024;24:75

15. Tetteh RA, et al. Prevalence and factors associated with syphilis among pregnant women in Ghana. BMC Pregnancy Childbirth. 2019;19:242

16. Eduku A, Senoo-Dogbey VE. Seroprevalence of hepatitis B virus infection (HBsAg) and associated factors among antenatal clinic attendees in a secondary-level facility in southern Ghana. Clin Epidemiol Glob Health. 2024;26:101553

17. World Health Organization. Hepatitis B. 2022. Available from: https://www.who.int/news-room/fact-sheets/detail/hepatitis-b

18. Agyare A, Amankwa AM, Owusu SK. Prevalence and associated factors of sexually transmitted infections among pregnant women in selected health facilities in Ghana. PLoS One. 2024;19(5):e0295088

19. Abesig J, Darko T, Ephraim RKD, Osakunor DNM, Nguah SB. Prevalence of viral hepatitis B among pregnant women in Ghana: a systematic review and meta-analysis. BMC Public Health. 2020;20(1):1–10

20. Ghana Health Service. National HIV and syphilis sentinel survey report. Accra: GHS; 2022

21. UNAIDS. UNAIDS data 2024. Geneva: Joint United Nations Programme on HIV/AIDS; 2024

22. Thompson K, et al. HIV and syphilis testing during pregnancy in Ghana. AIDS Res Hum Retroviruses. 2020;36(10):825–834

23. Owusu S, Adjei E, Boamah J. Urbanization and sexually transmitted infections among pregnant women in Ghana. BMC Public Health. 2021;21:324

24. Abbasi F, Niaz S, Irfan M, et al. Hepatitis C infection seroprevalence in pregnant women: a multicountry study. eClinicalMedicine. 2023;66:102875

25. Anabire NG, Wu L, Huang S, Wu L. Hepatitis C virus infection and risk factors among pregnant women in the Northern region of Ghana. Virol J. 2021;18(1):219

26. World Health Organization. Hepatitis C. 2022. Available from: https://www.who.int/news-room/fact-sheets/detail/hepatitis-c

27. Njau AF, Robert M, Rwebembera A, Kisendi CR, Maro H, Dennis G, et al. Prevalence and associated factors for HIV, HBV and syphilis coinfections among pregnant women attending antenatal care in Tanzania. PLoS One. 2025. doi:10.1371/journal.pone.0329068

28. Anteneh DE, Taye EB, Seyoum AT, Abuhay AE, Cherkose EA. Seroprevalence of HIV, HBV, and syphilis co-infections and associated factors among pregnant women in Amhara region, Ethiopia. PLoS One. 2024;19(8):e0308634

29. Dapaah J, et al. Epidemiology and determinants of hepatitis B and C infections in pregnant women attending antenatal clinics in Ghana. J Viral Hepat. 2024;31(2):164–172

30. Manyahi J, et al. Syphilis infection rates among pregnant women in Tanzania. BMC Infect Dis. 2015;15:501

31. Akinbami O, Rabiu KA, Adewunmi AA, Wright OJ, Dosunmu AO, Adewumi AA, Odum CU. Seroprevalence of syphilis, hepatitis B, C and HIV and their co-infections among pregnant women in Lagos, Nigeria. Ann Afr Med. 2013;12(3):182–187

32. Nankya I, et al. Burden of sexually transmitted infections among pregnant women in Uganda: a cross-sectional study. Trop Med Int Health. 2019;24(1):122–129

33. Baffoe P, Amoako YA, Ofosu W, Doku E, Helegbe GK. Determinants of syphilis and hepatitis B virus infections among pregnant women attending antenatal clinics in Ghana. Ghana Med J. 2022;56(1):34–41

34. Dionne-Odom JJ, et al. Prevalence and predictors of high-risk human papillomavirus infection among HIV-positive women in Malawi. Infect Agents Cancer. 2016;11:46

